# Specific Associations Between Type of Childhood Abuse and Elevated C-Reactive Protein in Young Adult Psychiatric Rehabilitation Participants

**DOI:** 10.1101/2022.04.04.22273426

**Authors:** Mbemba M. Jabbi, Philip D. Harvey, Raymond J. Kotwicki, Charles B. Nemeroff

## Abstract

**Background:** Early life adversity such as childhood emotional, physical, and sexual trauma is associated with a plethora of later-life psychiatric and chronic medical conditions, including elevated inflammatory markers. Although previous research suggests a role for chronic inflammatory dysfunctions in several disease etiologies, specific associations between childhood trauma types and later life inflammation and health status are not well understood.

**Methods:** We studied patients (n=280) who were admitted to a psychiatric rehabilitation center. Self-reported histories of childhood emotional, physical, and sexual trauma history were collected. At the time of admission, we also assessed individuals’ body mass index (BMI) and collected blood samples that were used to examine levels of inflammatory marker C-reactive protein (CRP).

**Results:** The prevalence of all three types of abuse were quite high, at 21% or more. 50% of the sample had elevations in CRP, with clinically significant elevations in 26%. We found that compared to a history of emotional or physical abuse, a history of childhood sexual trauma was more specifically associated with elevated CRP. This result held up when controlling for BMI.

**Limitation:** Our sample is relatively young, with an average age of 27.2 years, with minimal representation of ethnic and racial minority participants.

**Conclusion:** Relative to childhood emotional and physical trauma, childhood sexual trauma may lead to elevated inflammatory responses, which were common overall in the sample. Future studies need to assess the causal link between childhood sexual trauma and poorer health outcomes later in life.

**HIGHLIGHTS:**

- - The prevalence of both childhood abuse experiences and elevations in inflammatory markers were quite high.
- - We found that the history and severity of childhood sexual abuse were differentially correlated with later life inflammatory status and body mass index, with childhood emotional and physical abuse not showing the same degree of correlation with inflammatory status later in early adulthood.
- - These results demonstrate how specific elements of environmental adversity, which, when suffered at a critical developmental period, can have lingering negative physiological consequences later in life.

## INTRODUCTION

Childhood and adolescence are pivotal developmental periods during which physical and behavioral functions develop to shape later life adaptive functioning. Thus, traumatic environmental experiences during critical periods for a developing individual can have profound and lasting negative consequences on developmental and later-life health outcomes (Schrepf et al., 2014; Keicolt-Glaser et al., 2015; Petrov et al., 2016; Moraes et al., 2017; Jonker et al., 2017). For instance, previous research showed that childhood trauma (Gould et al., 2012), including emotional, physical, and sexual abuse or other forms of maltreatment is associated with cognitive impairments and poses long-lasting negative health consequences and significant morbidity and premature mortality (Edwards et al., 2012; Lu et al., 2013; Matthews et al., 2014; Pereira et al. 2019; Teicher et al., 2021; Lippard & Nemeroff 2020; Lippard & Nemeroff 2021).

Although research is beginning to identify the consequences of childhood adversity in terms of the negative impacts it exerts on central nervous system (CNS) and peripheral biomarkers, the precise linkage between childhood abuse history and later-life physical and psychological morbidity is not fully understood. Previous research revealed that over half of all developing children or their caregivers report at least one substantial traumatic experience before reaching 18 (Saunders & Adam 2014). Traumatic incidents of perceived or experienced violations of bodily integrity in the forms of emotional abuse, including the threat of physical harm, physical abuse, including actual injury resulting from violence, or the experience of sexual abuse, can have far-reaching negative consequences on later life emotional and physical well-being, morbidity, and poorer health outcomes (Scheffers M et al., 2017; Lippard and Nemeroff, 2020). However, whether these different childhood trauma subtypes and the severity of their occurrences can differentially influence later life mental and physical health outcomes is not well-understood.

Early-life trauma increases the risk of several psychiatric disorders, including depression (Kiecolt-Glaser et al., 2015), anxiety disorders, and PTSD (Choi & Sikkema 2016; Gould et al., 2021). Moreover, even in domains of severe mental illness, early life trauma appears to be associated with an increased risk for the development of schizophrenia among individuals equally predisposed by having a first-degree relative with the disease (Morgan & Anderson 2016). Further, individuals with early life trauma are more likely to develop treatment-resistant mood disorders and manifest multiple features of poor disease outcomes, including suicidal behaviors (Kiecolt-Glaser et al., 2015; Tunnard et al., 2014; Yrondi et al., 2021).

More recently, the effects of early life trauma on inflammatory responses and long-term dysregulations in immune functioning have been demonstrated (Pace et al., 2006; Kiecolt-Glaser et al., 2015; Pitharoulli et al., 2021; Danese and Lewis, 2017). For example, circulating C-reactive protein (CRP), an acute-phase protein of hepatic origin that rises in response to inflammation, can be increased after early life trauma (Keicolt-Glaser 2015; Pitharoulli et al., 2021). Elevated levels of CRP are also associated with cardiovascular disease (Kiecolt-Glaser 2015; Ananthan & Lyon 2020; Pitharoulli et al., 2021), obesity (Matthews et al., 2014), and certain cancers (Ananthan & Lyon 2020). In addition, elevated CRP concentrations (Keicolt-Glaser 2015; Ananthan & Lyon 2020; Pitharoulli et al., 2021) in traumatized individuals later in life can be associated with not only abuse histories but also with elevated measures of body mass index (BMI), anxiety, as well as depressive symptoms later in life (Noll et al., 2007).

The three types of early life abuse measured with the childhood trauma questionnaire (CTQ): physical, emotional, and sexual abuse (Bernstein et al., 1994; 1997 & 2003), may have different implications for later-life physical and mental health outcomes. For instance, in a study of newly admitted adolescent inpatients, a history of emotional abuse, but not physical or sexual abuse, correlated with being cyberbullied immediately before admission to an inpatient psychiatric unit (Saltz et al., 2020). Likewise, in a study of risk for developing PTSD in the immediate aftermath of trauma (Gould et al., 2021), the total severity of early life trauma, as measured by the total CTQ score, was associated with risk for the development of PTSD following this additional traumatic experience. In addition, many studies have suggested that sexual abuse is particularly pernicious in risk for later life psychopathology (Kiecolt-Glaser 2015; Baumeister et al., 2016; Ananthan & Lyon 2020; Pitharoulli et al., 2021).

Matthews and colleagues examined the relationship between CRP levels and childhood sexual, emotional, and physical abuse and neglect in a cohort of mid-life women (Matthews et al., 2014). They found that sexual and emotional abuse history, history of emotional and physical neglect, and the total number of recorded incidents of childhood abuse were associated with elevated CRP levels over a 7-year period. These findings of correlations between childhood abuse history and number of abuses with later life CRP levels were found to be moderated by BMI (Matthews et al., 2014). All participants were female, and all were outpatients. In addition, recent integrative reviews and meta-analyses examining the relationship between the history of childhood adversity and later life measures of CRP, including measures of interleukin family of inflammatory factors, and tumor necrosis factor-α (TNF-α), have found differential relationships between the type of trauma and later life inflammatory measures, but the results of these meta-analyses were not clear-cut (Shrepf et al. 2014; Baumeister et al. 2016; Pereira et al. 2019; Brown et al. 2021).

In the present study, we examined young (Mean age=27) females and males, varying in their lifetime trauma history, at the time of admission to a psychiatric rehabilitation facility. We assessed the presence and severity of early life trauma by documenting the history of a) emotional abuse, b) physical abuse, and c) sexual abuse using the CTQ (Bernstein et al., 1994; 1997 & 2003). Our study design affords a potentially reliable assessment of childhood trauma by assessing a young adult sample with limited chronicity of psychiatric and medical conditions and more proximal experiences of childhood trauma. Obtaining BMI and CRP data at the time of admission during the first physical examination, precluded treatment-related changes in CRP or BMI from influencing the results. These assessments at the time of admission allowed for the exclusion of individuals with chronic medical conditions that could cause elevations in inflammatory markers. We specifically examined the relationship between emotional, physical, and sexual trauma history (particularly meeting criteria for a definite history of abuse), and a) levels of CRP, and b) body mass index (BMI)/obesity. Previous studies have indicated that sexual abuse may be particularly pernicious both for later effects on functioning and with elevations in BMI (Moraes et al., 2017). In addition, previous studies in depression have highlighted the relationship between CRP levels and obesity (Pitharouli et al., 2021). Since elevated CRP and BMI have both been implicated as biophysical consequences of abuse (Matthews et al., 2014; Kiecolt-Glaser 2015; Ananthan & Lyon 2020; Pitharoulli et al., 2021; Noll et al., 2007), we examined the independent association of BMI and CRP with each of the three types of early-life abuse measured by the CTQ.

## METHODS

### Participants

This study comprised patients admitted to Skyland Trail, a non-profit residential and ambulatory psychiatric rehabilitation facility in Atlanta, GA. The center has a continuum of care in the context of a recovery model, where more symptomatic individuals were initially placed in residential facilities with planned transitions into day treatment, intensive outpatient, and transitional treatment tracks as symptomology improves. Therefore, less symptomatic individuals were directly entered into day treatment or intensive outpatient tracks. Individuals with chronic physical illnesses cannot be managed at Skyland Trail and are not admitted to residential services. Patients were included in the current analyses if they were admitted between April 2019 and December 2020 and completed the Childhood Trauma Questionnaire (CTQ), had a physical examination with a BMI score, and a blood sample collected at admission and assayed for CRP (n = 280). Diagnostic information was collected with the Mini-International Neuropsychiatric Inventory (MINI). Diagnoses using DSM-5 criteria were generated at admission with a consensus process involving a) trained clinicians who administered the structured rating scale and b) an attending psychiatrist who reviewed all available records and the clinician impressions. These diagnoses were collected at admission using a structured procedure on which we had previously published the methods (Kotwicki and Harvey, 2013).

The Institutional Review Board at the University of Miami Miller School of Medicine evaluated this project. That IRB determined that since all data were obtained for clinical purposes and were examined on a completely de-identified basis, the project is exempt from IRB review. See this determination in supplemental materials.

### Assessments

After admission and during their entire rehabilitation stay, patients at the center received biweekly clinical assessments tailored to their primary treatment targets. Participants with a diagnosis of major depression from the MINI interview were assessed with the Montgomery-Asberg Depression Rating Scale (MADRS) and the Hamilton Anxiety Rating Scale (HAM-A). Participants with a MINI diagnosis of bipolar disorder were also evaluated with the MADRS and the Young Mania Rating Scale (YMRS). The Brief Psychiatric Rating Scale (BPRS) was used to assess all cases with psychotic symptoms, including participants with major depression or bipolar disorder with psychotic features. Ratings were performed by raters who were not the primary clinicians. Thus, not all participants were rated with the same clinical assessment scales, for which we created an aggregate for use in statistical analyses as described below.

#### Childhood Trauma Questionnaire

The CTQ is a widely used 28-item self-report questionnaire that asks about experiences of abuse or neglect in childhood (Bernstein et al., 1994). It consists of five subscales measuring emotional, physical, or sexual abuse and emotional or physical neglect (Bernstein et al., 1994; 1997 & 2003). It has good internal consistency, and each subscale has items assessing minimization and denial, with all CTQ items reported on a 5 point scale ranging from Never True to Very Often True. The scale requires approximately a 6th-grade reading level to complete. We used the total scores for the CTQ and the three abuse subscales measuring a history of emotional, physical, or sexual abuse for our analyses. For our primary analyses, we generated present/absent ratings based on the criteria of Walker et al. (1999). These cutoffs include scores of 8 or more on sexual and physical abuse and 10 or more on emotional abuse.

#### C-Reactive Protein

Blood samples were collected at the time of admission during the physical examination from the antecubital vein, and CRP levels were measured at a commercial laboratory using the high sensitivity assay for CRP. For CRP, we used the total score and generated three different subclassifications of blood levels of CRP: <1.0 mg/L; 1.0 mg/L to 3.0 mg/L; and >3.0 mg/L. For reference, any CRP levels above the 0-3 mg/L are considered to reflect abnormally high/disease-related levels of peripheral inflammation (Pepys & Hirschfield 2003).

#### Body Mass Index (BMI)

We calculated BMI from height and weight measured during the admission physical examination when the blood sample for measurement of the CRP was also collected. As with the other variables, we collected BMI as a continuous variable and divided subjects into low or normal weight (BMI<24.9), overweight (BMI 25-29.9), and obese (BMI 30 or more) categories.

#### Data Analyses

Several analyses were performed. We examined the categorical associations between present absent ratings on the three CTA variables and the three classes of BMI and CRP measures. As a secondary analysis, we computed intercorrelations between the full range of raw scores on the three different abuse subscales and total scores on the CTQ and the full range of scores on BMI and CRP, using nonparametric Spearman Rank correlations. Finally, in the event of significant correlations between BMI and CRP and any of the categorical variables of abuse, we planned to use linear regression analysis to determine whether the associations between CRP and BMI and abuse were independent or if elevations in CRP were to some degree explained by increased body mass index. We also examined group differences in all four clinical rating scales with t-tests for the three abuse subtypes on a present/absent basis. We also created a composite clinical severity scale by standardizing all the scales and creating an aggregate score based on the average of the z scores for symptom severity for the individual scales.

We performed a power analysis to identify the size of correlations that could be detected meaningfully. Using the power analysis module of SPSS version 28 (IBM Corporation, 2021), we determined that a Pearson correlation of r=.17 could be detected with our sample size of 280, at p<.05, power=.80. Further, in terms of regression analyses, the power analysis suggested that we could detect an overall regression partial correlation of .18, at p<.05, power>.80. Thus, we had enough power to detect correlations that share a 3% variance between variables and would be at the lowest range of possible clinical significance.

## RESULTS

Descriptive information on the patient population is presented in **Table 1**. Participants were evenly divided between males and females (with an average age of 27.2) and the sample was largely white. Most were single and never married, and the principal diagnoses were in the mood disorders category. In addition, 46% had comorbid substance abuse, 42% were of average weight, and 28% were obese, with the high CRPs in the > 3.0 range being present in 26% of the participants.

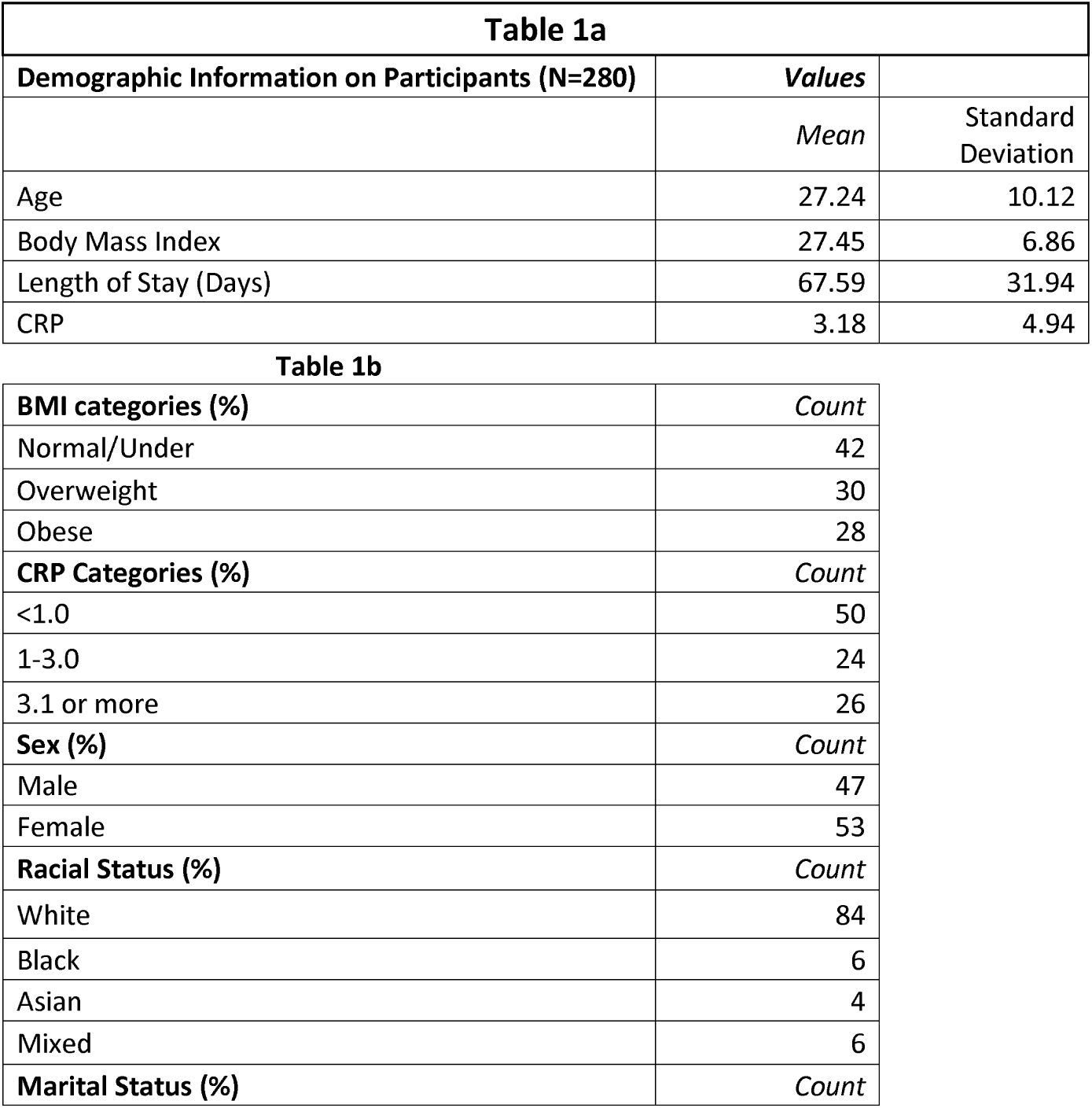
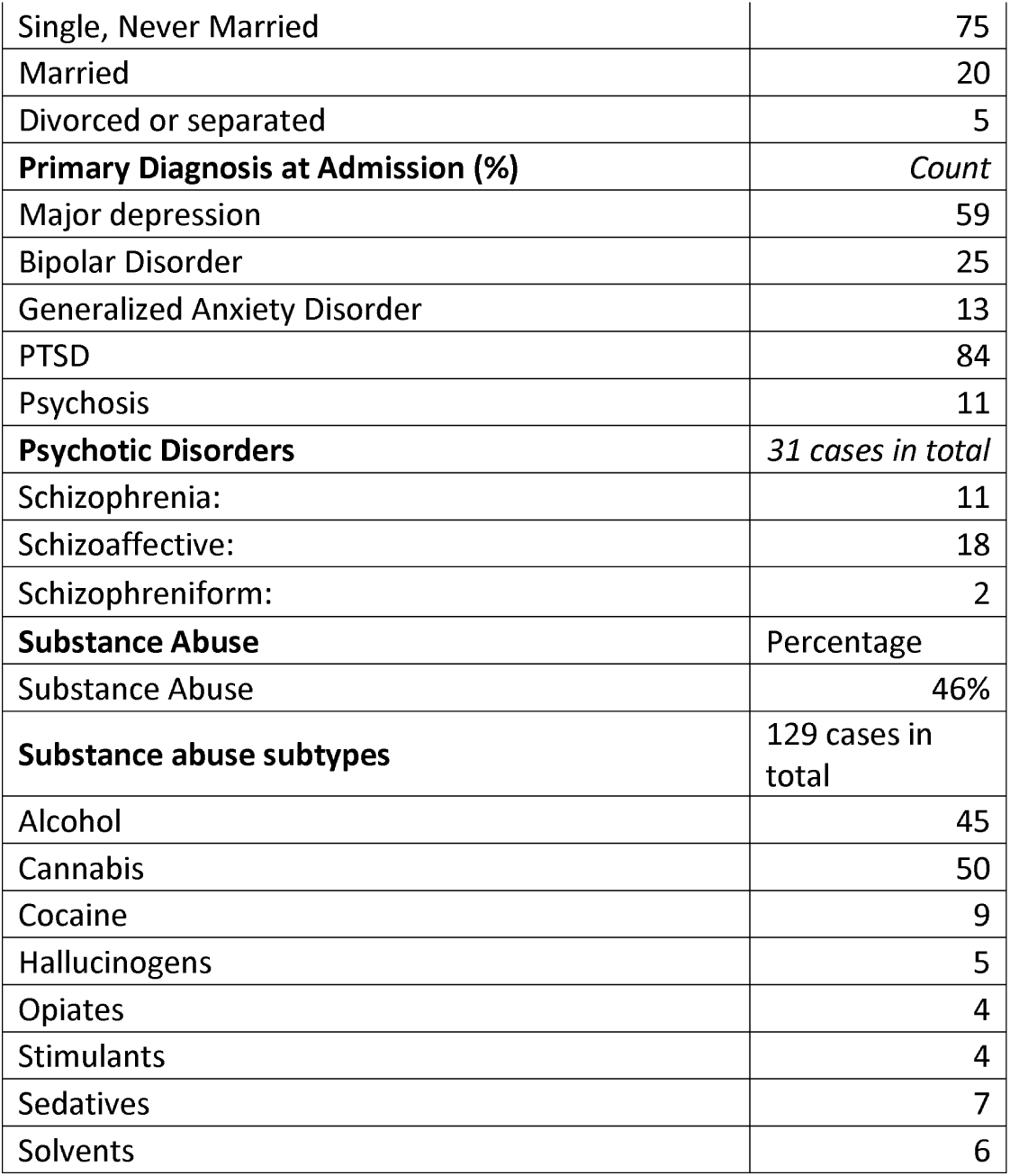

| **Table 1a** |  |  |
| --- | --- | --- |
| **Demographic Information on Participants (N=280)** | **Values** |  |
|  | *Mean* | Standard Deviation |
| Age | 27.24 | 10.12 |
| Body Mass Index | 27.45 | 6.86 |
| Length of Stay (Days) | 67.59 | 31.94 |
| CRP | 3.18 | 4.94 |

|  |  |
| --- | --- |
| **BMI categories (%)** | *Count* |
| Normal/Under | 42 |
| Overweight | 30 |
| Obese | 28 |
| **CRP Categories (%)** | *Count* |
| <1.0 | 50 |
| 1-3.0 | 24 |
| 3.1 or more | 26 |
| **Sex (%)** | *Count* |
| Male | 47 |
| Female | 53 |
| **Racial Status (%)** | *Count* |
| White | 84 |
| Black | 6 |
| Asian | 4 |
| Mixed | 6 |
| **Marital Status (%)** | *Count* |
| Single, Never Married | 75 |
| Married | 20 |
| Divorced or separated | 5 |
| **Primary Diagnosis at Admission (%)** | *Count* |
| Major depression | 59 |
| Bipolar Disorder | 25 |
| Generalized Anxiety Disorder | 13 |
| PTSD | 84 |
| Psychosis | 11 |
| **Psychotic Disorders** | *31 cases in total* |
| Schizophrenia: | 11 |
| Schizoaffective: | 18 |
| Schizophreniform: | 2 |
| **Substance Abuse** | Percentage |
| Substance Abuse | 46% |
| **Substance abuse subtypes** | 129 cases in total |
| Alcohol | 45 |
| Cannabis | 50 |
| Cocaine | 9 |
| Hallucinogens | 5 |
| Opiates | 4 |
| Stimulants | 4 |
| Sedatives | 7 |
| Solvents | 6 |

**Table 2** presents the means and standard deviations for the severity scores for abuse categories and the clinical variables and yes/no classifications based on our classification strategy; 53% were positive for emotional abuse, 27% for physical abuse, and 21% for sexual abuse. Furthermore, 27 of 280 cases (9.6%) were positive for all three forms of abuse. In comparison, both sexual abuse and emotional abuse (but not physical abuse) were positive for 17 of the 280 cases (6%), and both sexual abuse and physical abuse (but not emotional abuse) were present in only 2 cases (<1.0%).

**Table 2.** Childhood Trauma and Clinical Symptom Scores.

|  | **Sample**<br>**Size/N** | **Mean (% abuse history in sample)** | **Standard Deviation** |
| --- | --- | --- | --- |
| CTQ Total | 280 | 65.26 | 18.53 |
| Physical Abuse | 280 | 7.37 (27) | 4.53 |
| Emotional Abuse | 280 | 11.78 (53) | 6.33 |
| Sexual Abuse | 280 | 7.43 (21) | 5.75 |
| Montgomery-Asberg Depression | 187 | 22.78 | 8.63 |
| Hamilton Anxiety Rating Scale | 172 | 19.31 | 8.21 |
| Brief Psychiatric Rating Scale | 40 | 46.8 | 11.89 |
| Young Mania Rating Scale | 33 | 8.58 | 6.99 |

**Table 3** presents the categorical analyses of the abuse subtypes yes/no classification and BMI and CRP groupings. As seen in the table, for sexual abuse, but not emotional or physical abuse, there was a significantly higher number of participants with very high-level CRP scores, >3.0 mg/L. Obesity was not associated with the presence of any of the three types of abuse. We also examined other possible patient characteristics associated with the three different forms of abuse. We found that sex was not associated with any of the three abuse subtypes, all X^2^ (1)<1.91, all p>.39, and total scores on the CTQ did not differ as a function of sex, t(278)=.35, p=.73, and neither did length of stay did not correlate with CTQ total scores, ρ=-.07, p=.314. The occurrence of a substance abuse diagnosis did not differ as a function of the presence of any of the three types of abuse, all at X^2^ (1)<2.10, all p>.15.

**Table 3.**
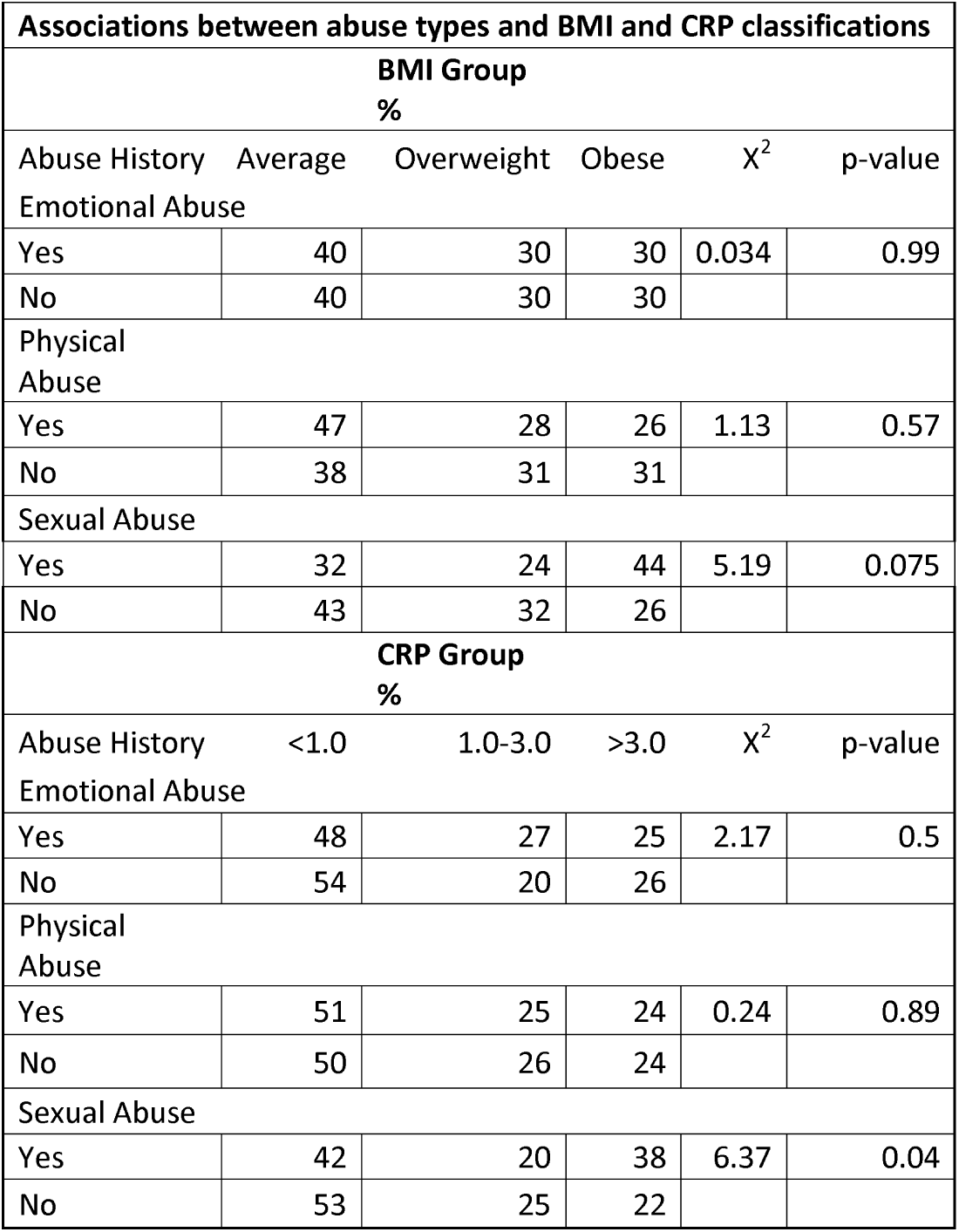

| **Table 3** |  |  |  |  |  |
| --- | --- | --- | --- | --- | --- |
| **Associations between abuse types and BMI and CRP classifications** |  |  |  |  |  |
| **BMI Group** |  |  |  |  |  |
| **%** |  |  |  |  |  |
| Abuse History | Average | Overweight | Obese | $\chi^2$ | p-value |
| **Emotional Abuse** |  |  |  |  |  |
| Yes | 40 | 30 | 30 | 0.034 | 0.99 |
| No | 40 | 30 | 30 |  |  |
| **Physical Abuse** |  |  |  |  |  |
| Yes | 47 | 28 | 26 | 1.13 | 0.57 |
| No | 38 | 31 | 31 |  |  |
| **Sexual Abuse** |  |  |  |  |  |
| Yes | 32 | 24 | 44 | 5.19 | 0.075 |
| No | 43 | 32 | 26 |  |  |
| **CRP Group** |  |  |  |  |  |
| **%** |  |  |  |  |  |
| Abuse History | <1.0 | 1.0-3.0 | >3.0 | $\chi^2$ | p-value |
| **Emotional Abuse** |  |  |  |  |  |
| Yes | 48 | 27 | 25 | 2.17 | 0.5 |
| No | 54 | 20 | 26 |  |  |
| **Physical Abuse** |  |  |  |  |  |
| Yes | 51 | 25 | 24 | 0.24 | 0.89 |
| No | 50 | 26 | 24 |  |  |
| **Sexual Abuse** |  |  |  |  |  |
| Yes | 42 | 20 | 38 | 6.37 | 0.04 |
| No | 53 | 25 | 22 |  |  |

**Table 4** presents intercorrelations between the CTQ variables and their correlations with BMI and CRP and group differences (Abuse present/Absent) in the four clinical variables. All abuse variables were significantly intercorrelated with Spearman’s rank correlation. We found associations between childhood abuse type and CRP to be significant only for sexual abuse severity-CRP correlations (ρ=.168, p=0.005), consistent with the analyses presented above regarding present/absent status for sexual abuse and being in the most elevated group for CRP. In contrast, and consistent with the group-based analyses, we did not find any significant correlations between CRP and emotional (ρ=0.038, p=.530) or physical (ρ=-.036, p=.549) abuse severity. BMI did not correlate with a history of emotional abuse (ρ=0.02, p=.252), physical abuse (ρ =0.02), or sexual abuse, ρ=0.13, p=.07. Furthermore, group differences based on presence/absence of childhood abuse subtype in relation to symptom ratings were observed only for Hamilton Anxiety scores, wherein physical and sexual abuse history, but not the history of emotional abuse, were associated with significantly higher baseline scores of anxiety. Depression, mania, and psychotic symptoms as indexed by the BPRS did not differ significantly between the different abuse groups, and the composite score of BPRS did not vary as a function of abuse history either. Finally, the presence of PTSD comorbidity (which was found in 84 of the 280 participants, with 58 comorbid for MDD and PTSD), was significantly associated with total CTQ scores. Those with comorbid PTSD had higher scores on the total CTQ (p=0.016, t(278)=2,42), as well as emotional (p<0.001,t(278)=4.12), physical (p=0.003, t(278)=3.03) and sexual abuse (p<0.001, t(278)=5.32). However, PTSD comorbidity was not related with either CRP levels (p=.47, t(278)=.49) or BMI (p=.12, t(278)=.73).

**Table 4.**

| **Table 4** |  |  |  |  |  |  |
| --- | --- | --- | --- | --- | --- | --- |
| **Associations of CTQ Scores, Clinical Variables, CRP, and Body Mass Index Based on Spearman Correlations** |  |  |  |  |  |  |
|  | **Emotional Abuse** | **Physical Abuse** | **Sexual Abuse** | **CRP** | **BMI** |  |
| CTQ Total | .59** | .83** | .77** | -0.02 | 0.04 |  |
| Emotional Abuse | -- | .66** | .40** | -0.06 | 0.02 |  |
| Physical Abuse | -- | -- | .56** | -0.07 | 0.04 |  |
| Sexual Abuse |  |  | -- | .17* | 0.13 |  |
| CRP |  |  |  | -- | .52** |  |
| Note. *p<.01; **p<.001 |  |  |  |  |  |  |
| **Group Differences in Clinical Variables as a Function of Abuse Status** |  |  |  |  |  |  |
|  | **Emotional Abuse** |  | **Physical Abuse** |  | **Sexual Abuse** |  |
| **Clinical Variable** | t-value | p-value | t-value | p-value | t-value | p-value |
| Montgomery-Asberg Depression | 0.91 | 0.36 | 1.46 | 0.07 | 0.21 | 0.82 |
| Hamilton Anxiety Rating Scale | 1.92 | 0.06 | 2.57 | 0.01 | 2.53 | 0.01 |
| Brief Psychiatric Rating Scale | 0.93 | 0.36 | 1.67 | 0.1 | 1.12 | 0.26 |
| Young Mania Rating Scale | 1.29 | 0.2 | 1.08 | 0.28 | 0.6 | 0.53 |
| Composite Score | 1.41 | 0.16 | 0.69 | 0.49 | 1.67 | 0.1 |

The severity of sexual abuse was associated with CRP on both dichotomous and continuous analyses, so, despite the lack of correlation between BMI and CRP, we used a regression analysis to evaluate whether the relationships between CRP, BMI, and sexual abuse were overlapping. We predicted CRP scores in the entire sample, entering BMI in the first block of regression analysis and then entering the three abuse severity scores in the second block, in a stepwise model. The overall regression analysis was significant at F(2,277)=10.80, p<.001. Specifically, the BMI scores entered first in the regression accounting for 5% of the variance in CRP scores (partial correlation: r=.22, t=2.75, p<.001), while the severity of sexual abuse accounted for an additional 3% of the variance (Partial correlation: r=.17, t=2.09, p=.031). Physical and emotional abuse history did not predict CRP levels after the entry of BMI and sexual abuse in the regression model: both t<1.51, p>.13.

## DISCUSSION

In this study, we examined the relationship between the childhood trauma subtypes and later-life peripheral inflammation as assessed with CRP levels and found a significant\influence of childhood sexual trauma on inflammatory markers. On a present/absent basis, 53%, 27%, and 21% of the participants reported childhood experiences of emotional, physical, and sexual abuse, respectively. These results underscore the high prevalence of early life trauma in individuals receiving treatment for psychiatric disorders. Furthermore, as this was a rehabilitation facility and all participants were previously treated without recovery, these individuals comprising the current study sample presented, with more refractory disease.

We found that higher levels of CRP correlated significantly with only the presence of a history of sexual abuse. This observed relationship between sexual abuse and CRP scores could not be explained by our measures of acute mood and psychotic symptoms, as the severity of depression, mania, and psychotic symptoms did not differ between childhood trauma subtypes. In addition, the severity of anxiety at the time of admission was also found to be elevated in participants with a physical or sexual abuse history, but not emotional abuse.

Of interest, the presence of a history of sexual abuse was not as strongly related to obesity or general mental illness diagnoses. Notably, except for childhood sexual abuse history, childhood abuse did not show a significant relationship, on either a continuous or dichotomous basis, with later life circulatory CRP levels. Our current finding aligns in part with a previous study showing a trend towards a statistically significant link between childhood sexual and emotional but not physical trauma abuse with indices of physiological health in a middle-aged female-only cohort (Matthews et al., 2014). Our findings also support another recent longitudinal study that followed children and their parents from when the children are ages 9-18 and then sampled their inflammatory CRP levels between ages 18-23, finding that a history of childhood sexual abuse and bullying victimization were the only two abuse types associated with later life elevated CRP levels (Lob et al. 2022). Our results also lend credence to the suggestion that the traumatic nature of childhood sexual abuse renders it a far-reaching negative risk factor for later life poor health outcomes (Matthews et al., 20214; Kiecolt-Glaser 2015; Baumeister et al., 2016; Scheffers M et al., 2017; Ananthan & Lyon 2020; Lippard & Nemeroff 2020; Pitharoulli et al., 2021). Together, our results suggest that, possibly more than emotional and physical abuse, childhood sexual trauma may have an enduring influence on later life inflammation.

Of the 280 patients whose data were examined, 84 had comorbid PTSD of which 58 were diagnosed with primary MDD, and 20 had bipolar disorder. It may seem surprising that primary diagnoses of PTSD were not more common in our sample given the pervasive history of childhood trauma, which was present in 65.26% of our participants. As noted above, childhood abuse combined with recent traumatic experiences appears to increase risk of the near-term development of PTSD. However, trauma exposure in childhood is also clearly linked to mood disorders (Lippard & Nemeroff 2022), especially MDD (Gordon, Nemeroff & Felitti 2020; Teicher, Gordon & Nemeroff 2021) and bipolar disorder (Gordon, Nemeroff & Felitti 2020; Teicher, Gordon & Nemeroff 2021), leading some to suggest that the field of psychiatry needs to include childhood trauma-induced disorder as a special diagnostic category (Gordon, Nemeroff & Felitti 2020; Teicher, Gordon & Nemeroff 2021), much like proposals for transdiagnostic consideration of suicidal ideation and behavior.

It is important to note that, although our findings are based on a sample of well-characterized psychiatric patients in a relatively controlled environment, the following limitations deserve mention. *First*, our sample size of 280 participants is not large, and minorities are under-represented, making our study sample not representative of the US population. The average sample age of 27.2 years is also relatively young. *Second*, seventy-five percent of the participants were either single or never married at the time of the study, which is not surprising given the relatively young age range of the sample. *Third*, psychiatric diagnoses were unevenly distributed, with major depression predominating and psychosis being relatively rare. *Fourth*, there were no sex differences in the prevalence of different types of abuse. Still, all participants were receiving treatment for persistent psychiatric morbidities, which might be associated with a lack of difference in the prevalence of the abuse types. Fifth, the CRP measures could be impacted by medication use. Still, because our currently reported CRP data were collected at baseline (i.e., at the time of admission), we could not fully control for prior medication. A lack of reliable data on medication use before admission means the possible effects of medication use before admission to the facility could not be accounted for. However, the specific correlation between CRP and only one of the three abuse subtypes argues against generic effects of medication treatment or other systemic illness because it is not clear how this bias could be related to a single type of early childhood abuse. *Fifth*, forty-six percent of our sample had a history of comorbid substance use even though none of these clinical variables, including sex, showed any significant relationship with CRP or BMI status. *Finally*, while our focus on CRP measures from peripheral blood has yielded important findings, the interpretations of our results showing selective inflammatory CRP correlates for sexual abuse are limited by the lack of interactive biomarkers such as brain imaging measures (Sheffield et al. 2013), or glucocorticoid metrics of the HPA-axis responses (Stein et al. 1997; Sapolsky et al. 2000), and transcription factor activity such as nuclear factor-κB (NF-κB) pathway responses (Pace et al. 2012). Considering our findings of an interaction between sexual abuse history and CRP levels, additional measures of glucocorticoid levels such as cortisol at admission could reveal further mechanistic insights given the role of glucocorticoids as transcription factors that mediate immune/inflammatory responses, and in influencing both carbohydrate and blood glucose metabolic functions during stressful and traumatic life events (Sapolsky et al. 2000).

Future studies need to recruit more representative samples across a wider age range spanning socioeconomic and racial groups, including younger individuals, where the abuse may have been even more proximal, and older participants to examine the lifelong persistence of these effects. Finally, the relationship between treatment response and measures of inflammation is an important issue that needs to be addressed with follow-up data, which is now being collected at this research site.

## Conclusion

The high prevalence rate of childhood trauma in this psychiatric population and the link between abuse history and later life negative consequences was supported by our findings of a specific association between childhood sexual abuse and early-adulthood inflammatory status. This finding is a compelling example of how childhood environmental adversity, at a critical developmental period, can have lingering negative physiological consequences. Future studies need to include more representative samples and further examine the mechanism by which early-life sexual trauma can have negative implications for physical health outcomes.

**Figure 1.**
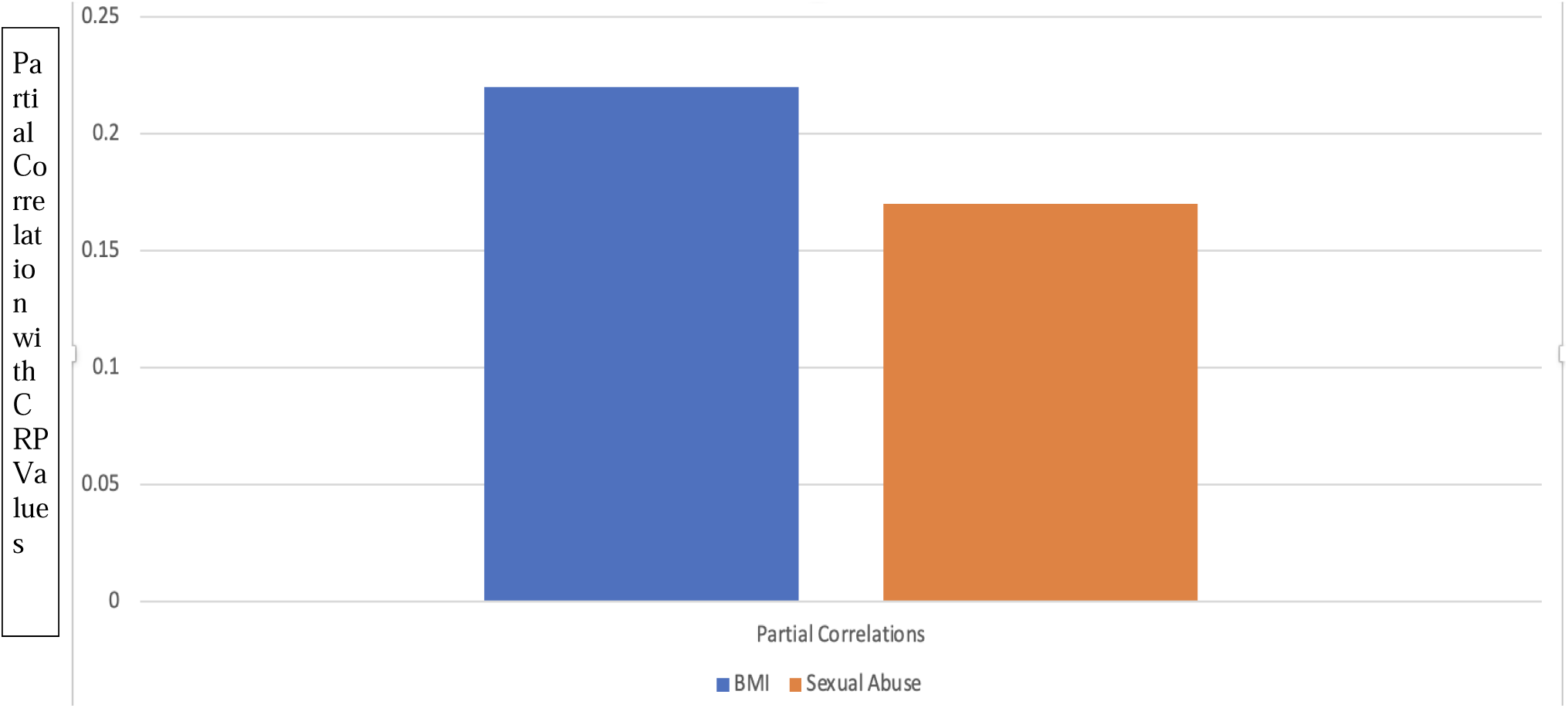
Illustrates the association between abuse history and BMI to predict CRP values across individuals. The stepwise regression indicated that only sexual abuse was correlated with CRP values.

## Data Availability

All data produced in the present study are available upon reasonable request to the authors.

## Acknowledgements

The authors thank the patients and family members for the data used in this study. The authors further thank Lokavya Marreddy for help with the preparation of this manuscript.

## Declaration of potential conflicts

**M Jabbi**.

None.

**PD Harvey** Dr. Harvey has received consulting fees or travel reimbursements from Alkermes, ANeuroTech (division of Anima BV), Bio Excel, Boehringer Ingelheim, Karuna Pharma, Minerva Pharma, and Sunovion Pharma during the past year. In addition, he receives royalties from the Brief Assessment of Cognition in Schizophrenia (Owned by Verasci, Inc). He is Chief Scientific Officer of i-Function, Inc.

**RJ. Kotwicki**.

None.

**CB Nemeroff**.

Research/Grants: National Institutes of Health (NIH)

Consulting (last 12 months): ANeuroTech (division of Anima BV), Signant Health, Sunovion Pharmaceuticals, Inc., Janssen Research & Development LLC, Magstim, Inc., Navitor Pharmaceuticals, Inc., Intra-Cellular Therapies, Inc., EMA Wellness, Acadia Pharmaceuticals, Axsome, Sage, BioXcel Therapeutics, Silo Pharma, XW Pharma, Neuritek, Engrail Therapeutics, Corcept Therapeutics Pharmaceuticals Company

Stockholder: Xhale, Seattle Genetics, Antares, BI Gen Holdings, Inc., Corcept Therapeutics Pharmaceuticals Company, EMA Wellness, TRUUST Neuroimaging

Scientific Advisory Boards: ANeuroTech (division of Anima BV), Brain and Behavior Research Foundation (BBRF), Anxiety and Depression Association of America (ADAA), Skyland Trail, Signant Health, Laureate Institute for Brain Research (LIBR), Inc., Magnolia CNS, Heading Health, TRUUST Neuroimaging

Board of Directors: Gratitude America, ADAA, Xhale Smart, Inc. Patents:

- Method and devices for transdermal delivery of lithium (US 6,375,990B1)
- Method of assessing antidepressant drug therapy via transport inhibition of monoamine neurotransmitters by ex vivo assay (US 7,148,027B2)

Speakers Bureau:

None

## Author Statement

Contributors: M Jabbi, P Harvey, R Kotwicki and C Nemeroff. P Harvey, R Kotwicki and C Nemeroff conceived and designed the study. R Kotwicki acquired the data. P Harvey and M Jabbi performed the data analysis. P Harvey, M Jabbi, R Kotwicki, and C Nemeroff wrote the paper. There was no external funding for this work.

